# Alpha globin gene copy number and hypertension risk among Black Americans

**DOI:** 10.1101/2022.02.02.22269915

**Authors:** A. Parker Ruhl, Neal Jeffries, Yu Yang, Orlando M. Gutierrez, Paul Muntner, Rakhi P. Naik, Lydia H. Pecker, Bryan T. Mott, Neil A. Zakai, Monika M. Safford, Leslie A. Lange, Cheryl A. Winkler, Marguerite R. Irvin, Mary Cushman, Hans C. Ackerman

**Author notes:** Corresponding authors: Dr. A. Parker Ruhl, MD, MHS, Building 10-Clinical Research Center, Rm. B3-4207, 10 Center Dr., Bethesda MD, 20892-1684, Hans Ackerman, MD, Dphil, 12735 Twinbrook Parkway, Room 3E-20, Rockville, Maryland 20852.

## Abstract

**BACKGROUND:** Alpha globin is expressed in the endothelial cells of human resistance arteries where it binds to endothelial nitric oxide synthase and limits release of the vasodilator nitric oxide. Genomic deletion of the alpha globin gene (*HBA*) is common among Black Americans and could lead to increased endothelial nitric oxide signaling and reduced risk of hypertension.

**METHODS:** Community-dwelling US adults aged 45 years or older were enrolled and examined from 2003 to 2007, followed by telephone every 6 months, and reexamined from 2013 to 2016. At both visits, trained personnel performed standardized, in-home blood pressure measurements and pill bottle review. Prevalent hypertension was defined as systolic blood pressure ≥ 140mmHg or diastolic blood pressure ≥ 90mmHg or anti-hypertensive medication use. Droplet digital PCR was used to determine *HBA* copy number. The associations of *HBA* copy number with prevalent hypertension, resistant hypertension, and incident hypertension were estimated using multivariable regression.

**RESULTS:** Among 9,684 Black participants, 7,439 (77%) had hypertension at baseline and 1,044 of those had treatment-resistant hypertension. 1,000 participants were not hypertensive at baseline and participated in a follow up visit; 517 (52%) developed hypertension over median 9.2 years follow-up. Increased *HBA* copy number was not associated with prevalent hypertension (PR=1.00; 95%CI 0.98,1.02), resistant hypertension (PR=0.95; 95%CI 0.86,1.05), or incident hypertension (RR=0.96; 95%CI 0.86,1.07).

**CONCLUSIONS:** There were no associations between increased *HBA* copy number and risk of hypertension. These findings suggest that variation in alpha globin gene copy number does not modify the risk of hypertension among Black American adults.

## INTRODUCTION

Black Americans have a higher prevalence of hypertension than other racial and ethnic groups such as White, Asian, and Hispanic Americans [1]. However, they and other minority racial and ethnic groups remain underrepresented in population studies of hypertensive disease [2,3]. A better understanding of genetic factors that influence blood pressure among Black Americans is needed to identify new targets for treatment of high blood pressure in this population.

The alpha subunit of hemoglobin is expressed in the endothelium of small arteries where it regulates the release of the vasodilator nitric oxide [4–6]. Specifically, alpha globin binds to endothelial nitric oxide synthase (eNOS) and reacts with nitric oxide to limit its diffusion into vascular smooth muscle. Silencing of the alpha globin gene, or targeting the alpha globin/eNOS complex pharmacologically, dilates isolated arteries and lowers blood pressure in mice [5,7,8]. In humans, alpha globin is expressed by the tandem duplicated genes *HBA1* and *HBA2* [9,10]. A common 3.7 kb deletion in the alpha globin gene cluster is common in people of African or Asian descent likely due to the protection it confers against severe malaria, [11–13] but how this deletion affects vasoregulation or vascular disease susceptibility is only beginning to emerge. The 3.7 kb alpha globin gene deletion is associated with enhanced dynamic vasodilation in adults of African or Asian ancestry [14,15]; however, the same deletion is not associated with blood pressure in healthy Kenyan adolescents [16]. We recently discovered this deletion to be associated with protection from chronic kidney disease and end-stage kidney disease in the Reasons for Geographic and Racial Differences in Stroke (REGARDS) study cohort, a large population-based cohort study of older Black adults [17]; however, the question of whether *HBA* copy number is associated with hypertension risk or blood pressure in this cohort remains unaddressed. To fill this knowledge gap, we analyzed the association of *HBA* copy number with prevalent hypertension, resistant hypertension, or incident hypertension in approximately 10,000 Black participants from the national longitudinal REGARDS cohort using multivariable models that included social, demographic, and biomedical risk factors for hypertension [18]. Among those not taking anti-hypertensive medications, we also examined the association of increasing *HBA* copy number with measured blood pressure.

## METHODS

### Study Design

REGARDS is a longitudinal cohort study designed to determine the reasons for racial disparities in stroke and cognitive decline in Black and White Americans aged ≥ 45 years.[18] REGARDS enrolled 30,239 participants from the 48 continental United States from 2003 to 2007. Exclusion criteria included self-reported race other than Black or White, residence in or on the waiting list for a nursing home, active cancer within the past year, or inability to communicate in English. A total of 12,514 (41%) REGARDS participants were Black, and 56% were from states considered to be the stroke belt where stroke incidence in the United States is highest. All self-reported Black participants consenting to genetic research were included in this study (Figure 1). All participants provided oral and written informed consent. The REGARDS study was approved by the Institutional Review Boards of the participating centers. This current study was determined to not be human subjects research by the NIH Office of Human Subjects Research Protection (OHSRP #13416) before this study began. This study followed the Strengthening the Reporting of Observational Studies in Epidemiology (STROBE) reporting guideline. The analytic plan was prespecified and approved by the REGARDS Cohort Study Executive Committee.

**Figure 1.**
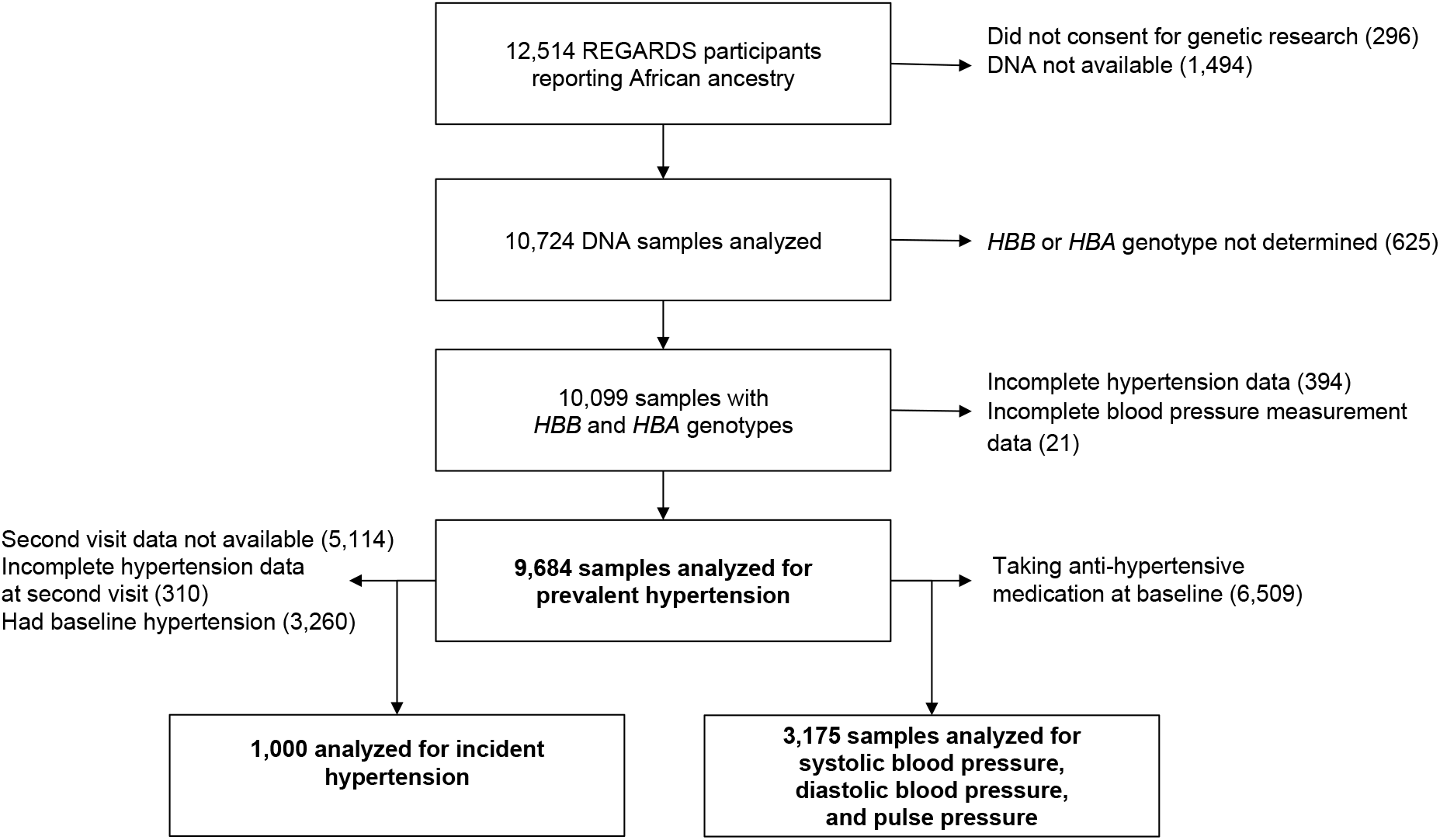
Study flow diagram REGARDS= REasons for Geographic and Racial Differences in Stroke Study; *HBB*= beta globin gene; *HBA*= alpha globin gene.

### Genotyping

We determined *HBA* copy number using droplet-digital polymerase chain reaction (ddPCR) analysis of genomic DNA as previously described [17]; the method is briefly described here and further details can be found in the online supplement. The assay targeted a sequence within the common 3.7 kb insertion/deletion polymorphism to provide a quantitative measure of alpha globin gene copy number per genome. Two-dimensional clusters of positive and negative droplets for the target and reference gene (*EIF2C1*) were manually gated using Quantasoft (Bio-Rad) per the manufacturer’s protocols. Droplet counts, copy number variant (CNV) values, and 95% CIs for CNV were extracted, visualized, and genotype was assigned using custom scripts in the R computing environment without user intervention. Copy number ranged from 2 to 6 gene copies, with 2 (-a/-a) and 3 (-a/aa) copies representing homozygosity and heterozygosity, respectively, for the 3.7 kb deletion, 4 (aa/aa) copies representing homozygosity for the reference allele, and 5 (aaa/aa) and 6 (aaa/aaa) copies representing heterozygosity and homozygosity for a known 3.7 kb insertion [19].

### Data collection

Baseline REGARDS study data were collected via standardized telephone interview, self-administered questionnaire, and an in-home visit. Trained personnel administered computer-assisted telephone interviews to collect study participant age, sex, region of residence, insurance status, education level, income, self-reports of prior physician-diagnosed comorbid conditions, and verbal informed consent. Trained personnel performed in-home examinations during which height, weight, and blood pressure were recorded, medication reviewed (with pill bottle review), electrocardiograms (EKG), and collection of blood and spot urine samples were performed, and written informed consent was obtained. These measurements were repeated in subjects who were alive and consented during a second in-home visit conducted a median (25^th^, 75^th^ percentile) of 9.5 (8.7, 9.9) years after their baseline study visit.

### Blood pressure measurement

Systolic blood pressure and diastolic blood pressure were measured by trained personnel using a standardized protocol recommended by the Joint National Committee on Prevention, Detection, and Treatment of High Blood Pressure (JNC 7) [20] with an aneroid sphygmomanometer (American Diagnostic Corporation, Hauppauge, NY) [18]. Participants were in a fasting state and seated for five minutes prior to blood pressure measurements.

Following a standardized protocol with an appropriate-sized cuff, an aneroid sphygmomanometer was used to measure systolic and diastolic blood pressure twice, with a 30-second rest period between the two measurements. The average of both measurements was used for all analyses.

### Outcome measures

Prevalent hypertension was defined as one or more of the following: 1) systolic blood pressure ≥ 140 mmHg or diastolic ≥ 90 mmHg; 2) self-reported current antihypertensive medication; or 3) two or more antihypertensive agents on in-home pill bottle review. Treatment-resistant hypertension was defined as taking medications from 4 or more antihypertensive classes or systolic blood pressure ≥ 140 mmHg or diastolic blood pressure ≥ 90 mmHg while taking medications from ≥ 3 antihypertensive classes. The number of classes of antihypertensive medication being taken at baseline was determined by pill bottle review during the baseline in-home study visit. Pulse pressure was defined as systolic blood pressure minus diastolic blood pressure. Incident hypertension was defined as systolic blood pressure ≥ 140 mmHg or diastolic blood pressure ≥ 90 mmHg or antihypertensive medication use at the follow-up home visit among those who did not have hypertension at baseline.

### Exposure variable

*HBA* copy number was evaluated as a numerical variable with values of 2, 3, 4, 5, or 6 as determined by ddPCR analysis of genomic DNA and modeled per increase of *HBA* copy number by one copy.

### Covariates

Age, sex, race, health insurance (yes or no), highest education level obtained (less than high school, high school, some college, college or more), annual income (≤ $20K, $20-34K, $35-74K, ≥ $75K), and smoking status (categorized by never, past, or current smoker) were self-reported.

Only those participants reporting Black race were included in the current analysis. Body mass index (BMI) was calculated from measured height and weight. Region was defined as three geographic areas: stroke belt buckle (coastal plains of North Carolina, South Carolina, and Georgia), stroke belt (the rest of North Carolina, South Carolina, and Georgia and the entire states of Tennessee, Alabama, Mississippi, Arkansas, and Louisiana), and stroke nonbelt (the remaining contiguous US) [21]. Chronic kidney disease was defined by the 2009 CKD-Epi equation and estimated glomerular filtration rate less than 60mL/min/1.73m^2^, urine albumin to creatinine ratio ≥ 30 mg/g measured on urine collected during the baseline in-home examination, or end-stage kidney disease. Fasting glucose levels ≥ 126 mg/dL, random glucose ≥ 200 mg/dL, or self-reported use of glucose-lowering medication was used to define diabetes mellitus. Total cholesterol was measured from blood collected during the in-home examination. C-reactive protein (CRP), hemoglobin, mean corpuscular volume (MCV), mean corpuscular hemoglobin (MCH), mean corpuscular hemoglobin concentration (MCHC), and red-cell distribution width-coefficient of variation (RDW-CV) values were measured or calculated from blood collected during the in-home examination [22]. CRP was natural log-transformed due to right skewness. The first 10 principal components of ancestry were calculated from Infinium Expanded Multi-Ethnic Genotyping Array data on 7,792 participants.

### Descriptive analysis of patient characteristics by HBA copy number

For continuous measures, median and 25^th^ and 75^th^ percentiles were reported by *HBA* copy number. Covariates and outcome measures were examined for outliers.

### Primary outcome

The primary outcome measure was prevalent hypertension. The association of *HBA* copy number with prevalent hypertension was evaluated first in an unadjusted regression model. A modified Poisson regression model with robust variance estimation for analyzing binary dependent variables was employed [23]. This Poisson approach gives the estimated risk ratio (RR) of hypertension for each covariate. The RR is regarded as a more clinically meaningful and transparent assessment of risk than the odds ratio in cohort studies [23]. The use of Poisson regression with robust variance estimation allows for estimating the RR directly and is reported here as the prevalence ratio (PR) for prevalent hypertension, rather than the odds ratio that arises from logistic regression. The covariates age, sex, BMI, region, insurance status, education level, income, hemoglobin, chronic kidney disease, diabetes mellitus, total cholesterol, and smoking status were included in all adjusted models. Additional models were estimated in pre-specified sensitivity analyses that included principal components of ancestry and CRP added separately to the base model and a stricter definition of resistant hypertension that required at least one antihypertensive medication to be a diuretic. Several post-hoc sensitivity analyses were performed. We removed chronic kidney disease and hemoglobin each separately from the base model. In addition, we evaluated the outcome of prevalent hypertension using the thresholds of systolic blood pressure ≥ 130 or diastolic blood pressure ≥ 80 mm Hg per the 2017 ACC/AHA recommendation [24].

### Secondary outcomes

The same modeling steps were used to analyze the following dichotomous outcomes: 1) the presence/absence of resistant hypertension at baseline reported as a PR, and 2) the presence/absence of incident hypertension (among those who had a second visit and were not hypertensive at baseline) reported as a RR (see Figure 1 illustrating participant follow up to the second visit). A similar set of models were estimated for the following continuous outcomes: systolic blood pressure, diastolic blood pressure, and pulse pressure. Multivariable linear regression was used to model these continuous outcomes. For these outcomes the *HBA* regression coefficients represent the estimated difference in the outcome measure associated with each additional *HBA* copy. The number of baseline anti-hypertensive medications in use was analyzed using a zero-inflated Poisson model [25,26]. This approach models the number of medications as a mixture of two latent, unobserved subpopulations: a ‘susceptible’ group that may have zero or more medications that is described by a Poisson distribution and a ‘non-susceptible’ group that has exactly zero medications. The non-susceptible population is included as part of the model because a Poisson model by itself has too few parameters to capture the relatively large number of individuals with zero medications (3175 of 9684). For this outcome the exponentiated *HBA* allele regression coefficient represents the multiplicative effect of an additional *HBA* allele on the expected number of medications among the susceptible subpopulation. Other exponentiated coefficients were similarly interpreted.

### Missing data considerations

Missing data for the primary outcome, secondary outcomes, and explanatory variables were typically rare (< 0.5%) with some exceptions, e.g., hemoglobin values (Table 1). Multiple imputation methods were employed in the multivariable models and methods as described in the online supplement [27,28].

**Table 1.** Clinical and demographic characteristics according to *HBA* copy number

|  | ***HBA* copy number** |  |  |  |  |  |
| --- | --- | --- | --- | --- | --- | --- |
| **Subjects, N (%)** | **All Subjects**<br>9,684 (100) | **2**<br>387 (4) | **3**<br>2685 (28) | **4**<br>6507 (67) | **≥ 5\***<br>105 (1) | ***P* value<sup>†</sup>**<br>NA |
| **Age, years** | 63<br>(57, 70) | 64<br>(58, 70) | 64<br>(57, 71) | 63<br>(57, 70) | 67<br>(61, 73) | 0.004 |
| **Female sex, N (%)** | 5952 (61) | 243 (63) | 1638 (61) | 4013 (62) | 58 (55) | 0.50 |
| **Body mass index, kg/m<sup>2</sup>** | 29.7<br>(26.1, 34.4) | 30.3<br>(26.9, 34.9) | 29.8<br>(26.1, 34.4) | 29.7<br>(26.1, 34.4) | 30.6<br>(27.5, 34.9) | 0.19 |
| **Smoking status, N (%)** |  |  |  |  |  | 0.58 |
| **Never** | 4368 (45) | 167 (43) | 1225 (46) | 2931 (45) | 45 (43) |  |
| **Past** | 3588 (37) | 156 (41) | 986 (37) | 2401 (37) | 45 (43) |  |
| **Current** | 1683 (17) | 61 (16) | 460 (17) | 1148 (18) | 14 (13) |  |
| **Region, N (%)** |  |  |  |  |  | 0.06 |
| **Non-Belt** | 4816 (50) | 169 (44) | 1367 (51) | 3220 (49) | 60 (57) |  |
| **Belt** | 3197 (33) | 138 (36) | 850 (32) | 2181 (34) | 28 (27) |  |
| **Buckle** | 1671 (17) | 80 (21) | 468 (17) | 1106 (17) | 17 (16) |  |
| **Medically insured, N (%)** | 8716 (90) | 350 (91) | 2428 (91) | 5842 (90) | 99 (91) | 0.61 |
| **Education level, N (%)** |  |  |  |  |  | 0.18 |
| **Less than high school** | 1896 (20) | 70 (18) | 532 (20) | 1271 (20) | 23 (21) |  |
| **High school graduate** | 2677 (28) | 111 (29) | 774 (29) | 1762 (27) | 30 (28) |  |
| **Some college** | 2572 (27) | 99 (27) | 738 (28) | 1707 (26) | 28 (28) |  |
| **College graduate or more** | 2527 (26) | 106 (27) | 639 (24) | 758 (27) | 24 (23) |  |
| **Income, N (%)** |  |  |  |  |  | 0.15 |
| **≤ \$20K** | 2600 (31) | 105 (30) | 721 (31) | 1742 (30) | 32 (33) | |
| **\$20K - \$34K** | 2513 (30) | 107 (31) | 711 (30) | 1681 (30) | 32 (33) | |
| **\$35K - \$74K** | 2489 (29) | 103 (30) | 708 (30) | 1651 (29) | 27 (28) | |
| **≥ \$75K** | 894 (10) | 32 (9) | 210 (9) | 670 (11) | 6 (6) | |
| **Chronic Kidney disease, N (%)** | 2498 (27) | 83 (22) | 688 (27) | 1692 (27) | 35 (35) | 0.05 |
| **Diabetes mellitus, N (%)** | 2818 (29) | 103 (27) | 808 (30) | 1878 (29) | 29 (28) | 0.47 |
| **Total Cholesterol, mg/dL** | 190<br>(165, 217) | 188<br>(162, 215) | 189<br>(164, 217) | 191<br>(165, 218) | 193<br>(172, 217) | 0.15 |
| **C-reactive protein, mg/L units** | 2.9<br>(1.2, 6.5) | 3.3<br>(1.3, 7.5) | 2.8<br>(1.1, 6.3) | 2.9<br>(1.2, 6.5) | 2.9<br>(1.1, 6.0) | 0.12 |
| **Hemoglobin, g/dL** | 13.1<br>(12.2, 14.0) | 12.4<br>(11.6, 13.2) | 12.9<br>(12.0, 13.8) | 13.2<br>(12.4, 14.1) | 13.0<br>(12.1, 14.1) | < 0.0001 |
| **MCV, fL** | 88<br>(84, 92) | 74<br>(72, 77) | 85<br>(82, 87) | 90<br>(87, 93) | 89<br>(87, 92) | < 0.0001 |
| **MCH, pg** | 29.6<br>(27.9, 30.9) | 23.8<br>(22.9, 24.8) | 27.9<br>(26.9, 29.0) | 30.3<br>(29.1, 31.4) | 29.9<br>(29.1, 31.1) | < 0.0001 |
| **MCHC, g/dL** | 33.4<br>(32.9, 33.9) | 32.1<br>(31.6, 32.6) | 33.0<br>(32.6, 33.5) | 33.6<br>(33.2, 34.1) | 33.8<br>(33.2, 34.0) | < 0.0001 |
| **RDW-CV, %** | 14.0<br>(13.3, 14.8) | 15.0<br>(14.4, 15.9) | 14.2<br>(13.5, 15.1) | 13.8<br>(13.2, 14.6) | 13.6<br>(13.2, 14.2) | < 0.0001 |
| **Prevalent hypertension<sup>‡</sup>, N (%)** | 7439 (77) | 304 (79) | 2070 (77) | 4978 (77) | 87 (83) | 0.20 |
| **Resistant hypertension, N (%)** | 1044 (11) | 50 (13) | 299 (11) | 679 (10) | 16 (15) | 0.16 |
| **Anti-hypertensive medication, N (%)** |  |  |  |  |  | 0.20 |
| **None** | 3175 (33) | 134 (35) | 877 (33) | 2135 (32) | 29 (28) |  |
| **One** | 1966 (20) | 67 (17) | 525 (20) | 1344 (21) | 30 (29) |  |
| **Two or more** | 4543 (47) | 186 (48) | 1283 (48) | 3028 (45) | 46 (44) |  |
| **Systolic blood pressure, mm Hg** | 130<br>(120, 140) | 130<br>(120, 140) | 130<br>(120, 141) | 130<br>(120, 140) | 133<br>(120, 144) | 0.42 |
| **Diastolic blood pressure, mm Hg** | 79<br>(72, 84) | 79<br>(71, 84) | 79<br>(72, 84) | 79<br>(71, 84) | 80<br>(71, 86) | 0.82 |
| **Pulse pressure, mm Hg** | 51<br>(43, 60) | 51<br>(43, 60) | 51<br>(43, 60) | 51<br>(43, 60) | 53<br>(45, 64) | 0.30 |

*HBA*= alpha globin gene; P= p value; N= number; K= thousand; RBC= red blood cell; MCV= mean corpuscular volume; MCH= mean corpuscular hemoglobin; MCHC= mean corpuscular hemoglobin concentration; RDW-CV= red cell distribution width-coefficient of variation; No.= number. Values are median (25<sup>th</sup>, 75<sup>th</sup> percentile) except where otherwise indicated.
\*105 subjects had 5 *HBA* gene copies and 3 subjects had 6 *HBA* copies; <sup>†</sup>P values for tests of differences by alpha globin copy number using chi-square test for categorical variables and Kruskal-Wallis test for continuous variables; <sup>‡</sup>261 participants met the definition of prevalent hypertension solely based on having two or more anti-hypertensive medications on pill bottle review.

### Data availability

The data underlying the findings include potentially identifying participant information and cannot be made publicly available because of ethical/legal restrictions. However, data including statistical code from this article are available to researchers who meet the criteria for access to confidential data. Data can be obtained upon request through the University of Alabama at Birmingham at. Additional information on the REGARDS study is available at www.regardsstudy.org.

## RESULTS

### HBA copy number variation and prevalent hypertension

*HBA* copy number was variable among the 9,684 Black study participants. 6,507 (67%) had four *HBA* copies, 2,685 (28%) had three copies, and 387 (4%) had two copies (Table 1). In addition, 105 (1%) participants had five *HBA* copies and three participants had six *HBA* copies, consistent with heterozygosity and homozygosity for the known *HBA* gene triplication.[19] Among the 9,684 participants, 7,439 (77%) had hypertension at baseline, 1,044 (11%) had resistant hypertension, and 4,543 (47%) were taking two or more antihypertensive medications (Table 1). In multivariable regression analyses, *HBA* copy number was not associated with prevalent hypertension, (PR = 1.00; 95%CI 0.98, 1.02), resistant hypertension, or the number of antihypertensive medications being taken (Table 2).

**Table 2.**
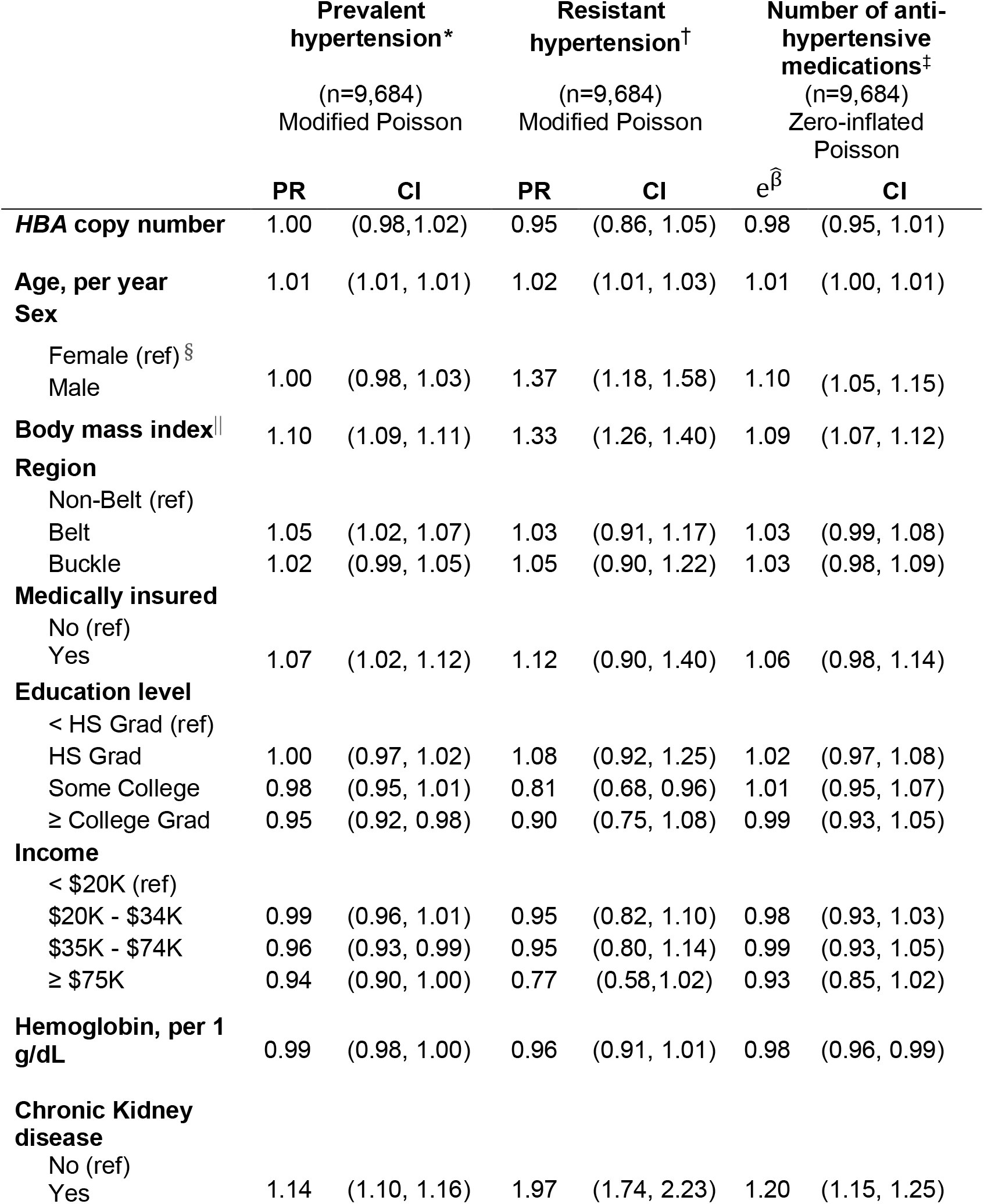
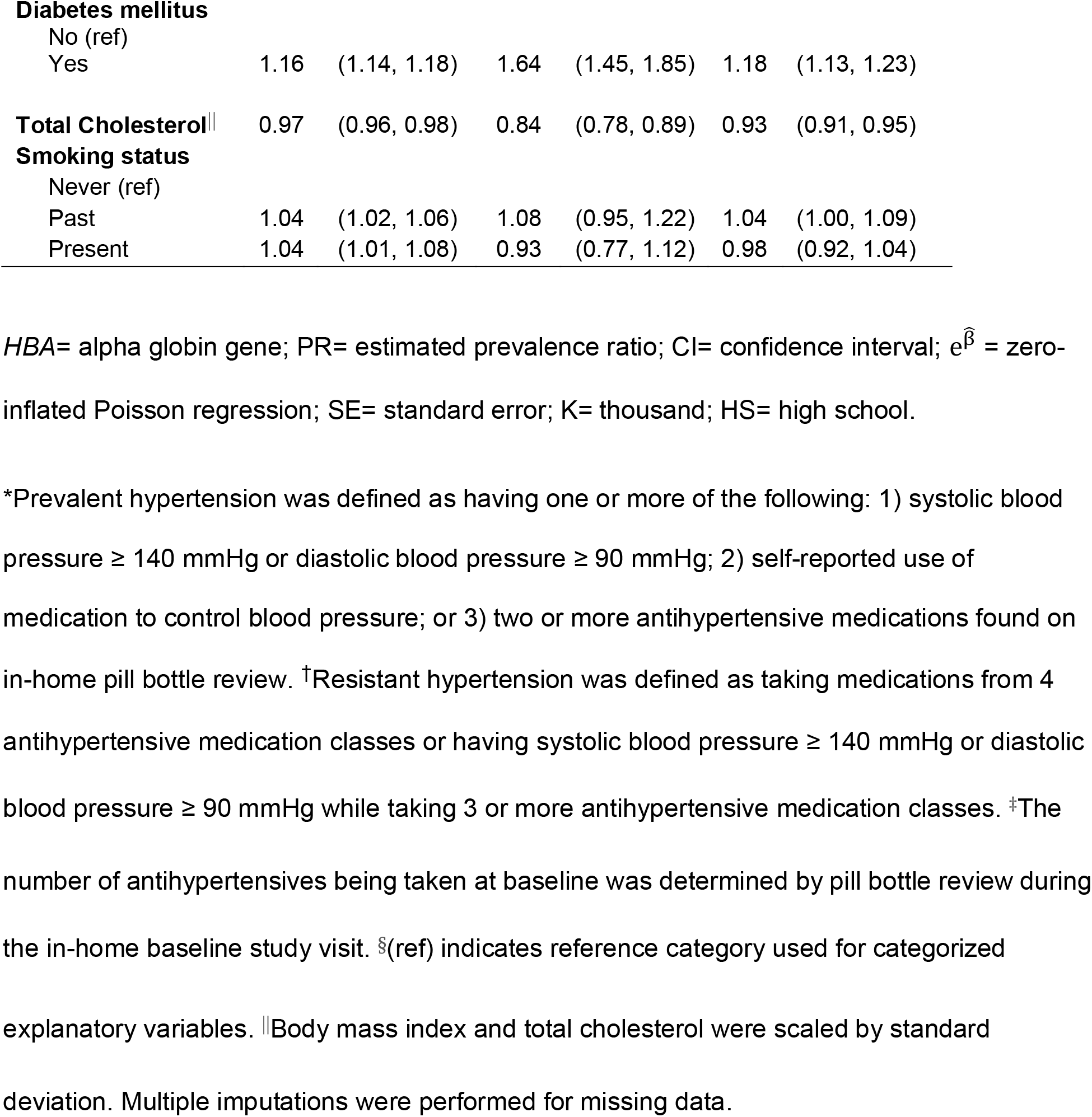
Association of *HBA* copy number with prevalent hypertension, resistant hypertension, or anti-hypertensive use–fully adjusted analyses

Missing data are as follows: medically insured (n=12, <1%); education (n=12, <1%); income (n=1,170 refused, 12%); kidney disease (n=389, 4%); diabetes mellitus (n=89, <1%); total cholesterol (n=57, <1%); body mass index (n=55, <1%); smoking status (n=45, <1%); c-reactive protein (n=151, 1.6%); hemoglobin (n=3,089, 32%); MCV (n=3,095, 32%); MCH (n=3,089, 32%); MCHC (n=3,089, 32%); RDW-CV (n=3,102, 32%). All other variables in Table 1 had no missing values.

### HBA copy number variation and incident hypertension

Among the 1,000 participants who were not hypertensive at baseline and who participated in the second visit, 517 (52%) developed hypertension over a median (25^th^, 75^th^ percentile) of 9.2 (8.6, 9.9) years of follow-up. In multivariable regression analysis, *HBA* copy number was not associated with the risk of incident hypertension (RR = 0.96; 95%CI 0.86, 1.07; Table 3).

**Table 3.** Association of *HBA* copy number with incident hypertension – fully adjusted analysis

|  | **Incident**<br>**Hypertension\*** |  |
| --- | --- | --- |
|  | (n=1,000)<br>Modified Poisson |  |
|  | **RR** | **CI** |
| ***HBA* Copy Number** | 0.96 | (0.86, 1.07) |
| **Age, per year** | 1.01 | (1.00, 1.02) |
| **Sex** |  |  |
| Female (ref) <sup>†</sup> |  |  |
| Male | 0.95 | (0.82, 1.11) |
| **Body mass index<sup>†</sup>** | 1.14 | (1.08, 1.20) |
| **Region** |  |  |
| Non-Belt (ref) |  |  |
| Belt | 1.12 | (0.98, 1.27) |
| Buckle | 0.99 | (0.84, 1.16) |
| **Medically insured** |  |  |
| No (ref) |  |  |
| Yes | 0.97 | (0.80, 1.17) |
| **Education level** |  |  |
| < HS Grad (ref) |  |  |
| HS Grad | 1.00 | (0.82, 1.23) |
| Some College | 0.95 | (0.77, 1.17) |
| ≥ College Grad | 0.83 | (0.67, 1.03) |
| **Income** |  |  |
| < \$20K (ref) | | |
| \$20K - \$34K | 1.01 | (0.84, 1.22) |
| \$35K - \$74K | 1.04 | (0.87, 1.25) |
| ≥ \$75K | 0.92 | (0.72, 1.18) |
| **Hemoglobin, per 1 g/dL** | 1.00 | (0.94, 1.06) |
| **Chronic Kidney disease** |  |  |
| No (ref) |  |  |
| Yes | 1.10 | (0.91, 1.32) |
| **Diabetes mellitus** |  |  |
| No (ref) |  |  |
| Yes | 1.30 | (1.12, 1.51) |
| **Total Cholesterol<sup>‡</sup>** |  |  |
|  | 0.94 | (0.88, 1.01) |
| **Smoking status** |  |  |
| Never (ref) |  |  |
| Past | 0.98 | (0.85, 1.12) |
| Present | 1.15 | (0.97, 1.36) |

*HBA*= alpha globin gene; RR= risk ratio; CI= confidence interval; K= thousand; HS= high school.
\*Incident hypertension was defined as systolic blood pressure $\geq 140$ mmHg or diastolic blood pressure $\geq 90$ mmHg or reported medication use to control blood pressure during the REGARDS study cohort follow-up visit, among those who were not hypertensive at baseline (systolic blood pressure $< 140$ mmHg or diastolic blood pressure $< 90$ mmHg or no medication use to control blood pressure). The incident hypertension analysis was performed on 1,000 participants who were not hypertensive at baseline and had second visit data available. The median (25<sup>th</sup>, 75<sup>th</sup> percentile) time elapsed between the baseline and second visit was (9.2 [8.6, 9.9]) years. P values calculated by modified Poisson regression model for dichotomous outcome measures. <sup>†</sup>(ref) indicates reference category used for categorized explanatory variables. <sup>‡</sup>Body mass index and total cholesterol were scaled by standard deviation. Multiple imputations were performed for missing data.

### HBA copy number variation and measured blood pressure

The association between *HBA* copy number and measured blood pressure was assessed in 3,175 participants who were not taking antihypertensive medications at baseline. Each additional *HBA* copy was associated with lower diastolic blood pressure (−0.66 mmHg; 95%CI -1.25, -0.07; Table 4) but was not significantly associated with lower systolic blood pressure (−0.88 mmHg; 95%CI -1.89, 0.13; Table 4). There was no association between *HBA* copy number and pulse pressure (−0.22 mmHg; 95%CI -0.95, 0.52) as the direction of the effect of HBA copy number on systolic blood pressure and diastolic blood pressure was the same.

**Table 4.** Association of *HBA* copy number with measured blood pressure in subjects not taking anti-hypertensive medications–fully adjusted analyses

|  | Systolic blood pressure*<br>(n=3,175)<br>Multivariable linear |  | Diastolic blood pressure*<br>(n=3,175)<br>Multivariable linear |  | Pulse pressure*<sup>†</sup><br>(n=3,175)<br>Multivariable linear |  |
| --- | --- | --- | --- | --- | --- | --- |
| | $\hat{\beta}$ | CI | $\hat{\beta}$ | CI | $\hat{\beta}$ | CI |
| ***HBA* copy number** | -0.88 | (-1.89, 0.13) | -0.66 | (-1.25, -0.07) | -0.22 | (-0.95, 0.52) |
| **Age, by year** | 0.27 | (0.20, 0.34) | -0.06 | (-0.10, -0.02) | 0.33 | (0.28, 0.39) |
| **Sex** |  |  |  |  |  |  |
| Female (ref) |  |  |  |  |  |  |
| Male | 4.53 | (3.06, 5.99) | 2.18 | (1.33, 3.03) | 2.34 | (1.28, 3.40) |
| **Body mass index<sup>§</sup>** | 3.27 | (2.67, 3.88) | 2.23 | (1.87, 2.58) | 1.04 | (0.60, 1.48) |
| **Region** |  |  |  |  |  |  |
| Non-Belt (ref) |  |  |  |  |  |  |
| Belt | 0.40 | (-0.89, 1.69) | -0.10 | (-0.86, 0.65) | 0.50 | (-0.44, 1.44) |
| Buckle | -2.24 | (-3.84, -0.63) | -0.78 | (1.72, 0.15) | -1.45 | (-2.62, -0.28) |
| **Medically insured** |  |  |  |  |  |  |
| No (ref) |  |  |  |  |  |  |
| Yes | -1.83 | (-3.61, -0.05) | -0.53 | (-1.57, 0.51) | -1.30 | (-2.61, -0.28) |
| **Education level** |  |  |  |  |  |  |
| < HS Grad (ref) |  |  |  |  |  |  |
| HS Grad | -3.10 | (-4.99, -1.21) | -1.42 | (-2.53, -0.32) | -1.68 | (-3.06, -0.29) |
| Some College | -3.30 | (-5.23, -1.38) | -1.05 | (-2.18, 0.08) | -2.25 | (-3.66, -0.85) |
| ≥ College Grad | -4.17 | (-6.19, -2.15) | -1.56 | (-2.74, -0.37) | -2.61 | (-4.09, -1.13) |
| **Income** |  |  |  |  |  |  |
| < \$20K (ref) | | | | | | |
| \$20K - \$34K | 1.06 | (-0.63, 2.76) | 0.23 | (-0.76, 1.22) | 0.83 | (-0.42, 2.08) |
| \$35K - \$74K | -0.18 | (-1.96, 1.59) | -0.37 | (-1.42, 0.68) | 0.18 | (-1.13, 1.49) |
| ≥ \$75K | -0.99 | (-3.40, 1.42) | -0.65 | (-2.04, 0.73) | -0.34 | (-2.11, 1.44) |
| **Hemoglobin, per 1 g/dL** |  |  |  |  |  |  |
|  | -0.15 | (-0.70, 0.40) | 0.62 | (0.30, 0.94) | -0.77 | (-1.17, -0.38) |
| **Chronic Kidney disease** |  |  |  |  |  |  |
| No (ref) |  |  |  |  |  |  |
| Yes | 6.49 | (4.78, 8.19) | 2.96 | (1.95, 3.96) | 3.53 | (2.30, 4.76) |
| **Diabetes mellitus** |  |  |  |  |  |  |
| No (ref) |  |  |  |  |  |  |
| Yes | 0.58 | (-1.12, 2.27) | -0.92 | (-1.91, 0.08) | 1.50 | (0.25, 2.74) |
| **Total Cholesterol<sup>§</sup>** | 1.34 | (0.75, 1.92) | 0.65 | (0.30, 0.99) | 0.69 | (0.27, 1.12) |
| **Smoking status** |  |  |  |  |  |  |
| Never (ref) |  |  |  |  |  |  |
| Past | 0.27 | (-1.07, 1.60) | -0.72 | (-1.50, 0.06) | 0.98 | (-0.01, 1.96) |
| Present | 3.06 | (1.48, 4.64) | 0.52 | (-0.41, 1.45) | 2.54 | (1.39, 3.70) |

*HBA*= alpha globin gene; $\hat{\beta}$ = estimated linear regression coefficient; CI= confidence interval; K= thousand; HS= high school.
\*The analyses of measured blood pressured outcomes were performed only on those subjects not currently taking antihypertensive medication (n=3,175). <sup>†</sup>Pulse pressure was defined as systolic blood pressure minus diastolic blood pressure. <sup>‡</sup>(ref) indicates reference category used for categorized explanatory variables. <sup>§</sup>Body mass index and total cholesterol were scaled by standard deviation. Multiple imputations were performed for missing data

### Sensitivity and interaction analyses

Pre-specified sensitivity analyses were performed to address possible population stratification, to explore the role of inflammation in the relationship between *HBA* copy number and prevalent hypertension, and to evaluate a stricter definition of resistant hypertension. The sensitivity analysis for population stratification was performed by adding the first 10 principal components of ancestry to the prevalent hypertension model; this did not change the association of *HBA* copy number with prevalent hypertension (Table S1). To explore the role of inflammation, c-reactive protein (CRP) was included as a covariate in the multivariable models. Log CRP was associated with increased risk of prevalent hypertension (PR = 1.03; 95% CI 1.02,1.04) but did not change the association of *HBA* copy number with prevalent hypertension (Table S2). We evaluated an alternative definition of resistant hypertension that required at least one of the anti-hypertensive medications to be a diuretic, and there was no change in the association of *HBA* copy number with resistant hypertension (Table S3). We conducted pre-specified tests for statistical interaction between age, sex, or chronic kidney disease with *HBA* copy number on the outcome of prevalent hypertension and found no significant interactions (all p values for interaction terms >0.20, Table S4). Post-hoc sensitivity analyses removing hemoglobin or chronic kidney disease from the model did not change the association of *HBA* copy number with prevalent hypertension (Tables S5 and S6). Defining hypertension with a lower blood pressure threshold also did not change the association of *HBA* copy number with prevalent hypertension (Table S7).

## DISCUSSION

In light of the novel role of endothelial alpha globin as a restrictor of nitric oxide signaling in resistance arteries, we hypothesized that individuals who inherit a deletion of the alpha globin gene would be less likely to develop hypertension. To test this hypothesis, we used digital droplet PCR to genotype a 3.7 kb insertion/deletion polymorphism that quantitatively alters alpha globin gene copy number in a large longitudinal cohort of Black Americans. We found no association between *HBA* copy number and the pre-specified primary outcome of prevalent hypertension. There was also no association between *HBA* copy number and the secondary outcomes of prevalent resistant hypertension or the number of anti-hypertensive medications being taken. *HBA* copy number was not associated with incident hypertension over a decade of observation.

There have been no studies reporting the association between *HBA* copy number and the clinical outcome of hypertension for direct comparison. A related study of Kenyan adolescents found no association between the 3.7 kb gene deletion and 24-hour ambulatory blood pressure [16]. Our study extends that observation into a population of older Black American adults with a range of social, economic, and medical risk factors where we find no association between *HBA* copy number and the clinical outcome of hypertension. One relevant co-morbidity is chronic kidney disease, which was recently found to be associated with *HBA* copy number in a separate study also performed in the REGARDS cohort [29]. Chronic kidney disease was included in the pre-specified model in the current study evaluating the association of *HBA* copy number with hypertension. We performed a pre-specified test of interaction with *HBA* copy number and chronic kidney disease (Table S4) and a post-hoc sensitivity analysis removing chronic kidney disease from the main model (Table S6) and neither demonstrated any impact on the finding that there was no association between *HBA* copy number and hypertension. Similarly, removing hemoglobin from the main model did not alter the negative findings of this study (Table S5).

In contrast to the study in Kenyan adolescents where there was no association between *HBA* and blood pressure, we observed that each additional copy of *HBA* was associated with lower diastolic blood pressure among adults. This observation implies that increased expression of alpha globin is associated with lower blood pressure, rather than higher blood pressure as hypothesized. Keller et al. recently demonstrated that endothelial alpha globin can produce nitric oxide from the chemical reduction of nitrite under hypoxic conditions [30]. Thus, endothelial alpha globin may function not only as a nitric oxide scavenger, but also as a nitric oxide producer. Since NOS-dependent vascular nitric oxide production declines with age, the relative roles of alpha globin as a nitric oxide scavenger versus a nitric oxide producer may shift with age as well, potentially explaining the unexpected association of higher *HBA* copy number with lower diastolic blood pressure in this study of older Black Americans.

This study has strengths and limitations. The REGARDS Study is one of the largest cohorts of Black Americans, and the participants were well characterized using standardized measurements for blood pressure [20] and surveyed using instruments validated for collecting social and biomedical factors known to impact hypertension [31]. We used a robust and quantitative ddPCR approach to genotype the *HBA* insertion/deletion directly and avoided the potential for mis-assignment that can result from inferring genotype from sequence analysis or SNP genotyping. We focused on the most prevalent structural variant in Black Americans, the 3.7 kb deletion, but did not evaluate rare deletions or regulatory polymorphisms that could affect gene expression [32]. We utilized a pre-specified analysis plan to reduce the chance of identifying spurious associations, however, no statistical corrections were made for multiple testing.

While this study has strengths from an epidemiological perspective, the hypothesis tested relies on several implicit assumptions linking gene expression in the vascular endothelium to the phenotypes of blood pressure and hypertension. First, while *HBA* copy number is quantitatively associated with alpha globin expression in erythrocyte progenitors, the same relationship has not yet been established for endothelial cells in the wall of the resistance artery. In the future, measuring gene expression directly in human arteries would establish the relationship between *HBA* copy number and gene expression in this important cellular context. Second, arteries may adapt to the loss of endothelial alpha globin by downregulating the expression or activity of nitric oxide synthase or its downstream receptors such as soluble guanylate cyclase. Thus, compensation may obscure the effect of reduced endothelial alpha globin gene expression on blood pressure. Third, while deletion of *HBA* has been associated with enhanced dynamic vasodilation [14,15], blood pressure is regulated by multiple pathways that are integrated across the renal, cardiovascular, and nervous systems. Thus changes in arterial nitric oxide signaling could be overcome by other regulatory systems to maintain a blood pressure set point [33,34]. Moreover, endothelial nitric oxide synthase has a diminishing role in the regulation of vasodilation as people age and specifically in hypertension [35–39]. In the older population studied here, endothelial nitric oxide synthase may no longer be as active and thus would not be subject to regulation by different levels of alpha globin expression. Therefore, the absence of association between *HBA* copy number and the clinical phenotype of hypertension does not exclude a role for alpha globin as a regulator of nitric oxide signaling in human resistance arteries [4,6–8]. Further investigation into the role of alpha globin in the dynamic vasoreactivity of human resistance arteries remains warranted.

## PERSPECTIVES

Despite previous evidence that alpha globin regulates nitric oxide signaling in resistance arteries, we found no association between *HBA* copy number and risk of hypertension among older Black American adults.

## ACKNOWLEDGEMENTS

The authors thank the investigators, staff, and participants of the REGARDS study for their valuable contributions. A full list of participating REGARDS investigators and institutions can be found at http://www.regardsstudy.org.

