## Supplement for "Alpha globin gene copy number and hypertension risk among Black Americans"

[1]Laboratory of Malaria and Vector Research, National Institute of Allergy and Infectious Diseases, National Institutes of Health, Bethesda, Maryland; [2]Pulmonary Branch, National Heart, Lung, and Blood Institute, Bethesda, Maryland; [3]Office of Biostatistics Research, National Heart, Lung, and Blood Institute, Bethesda, Maryland; [4]Division of Blood Diseases and Resources, National Heart, Lung, and Blood Institute, Rockville, Maryland; [5]Department of Medicine, University of Alabama at Birmingham, Birmingham, Alabama, [6]Department of Epidemiology, University of Alabama at Birmingham, Birmingham, Alabama [7]Division of Hematology, Department of Medicine, Johns Hopkins University School of Medicine, Baltimore, Maryland; [8]University of Alabama at Birmingham School of Medicine, Birmingham, Alabama; [9]Department of Medicine, Larner College of Medicine at the University of Vermont, Burlington, Vermont; [10]Department of Pathology & Laboratory Medicine, Larner College of Medicine at the University of Vermont, Burlington, Vermont; [11]Department of Medicine, Weill Cornell Medicine, New York, New York; [12]Division of Biomedical Informatics and Personalized Medicine, Department of Medicine, University of Colorado, Denver, CO; Department of Epidemiology, [13]Basic Research Laboratory, National Cancer Institute, Frederick National Laboratory for Cancer Research, Frederick, Maryland

Table of Contents

I. Supplemental Tables
   a. Table S1. Sensitivity analysis for the association of *HBA* copy number with prevalent hypertension – fully adjusted model with the addition of the principal components of ancestry

   b. Table S2. Sensitivity analysis for the association of *HBA* copy number with prevalent hypertension – fully adjusted model with the addition C-reactive protein

   c. Table S3. Sensitivity analysis for the association of *HBA* copy number with resistant hypertension requiring diuretic use – fully adjusted model

   d. Table S4. Pre-specified tests for interaction between *HBA* copy number and age, sex, and chronic kidney disease on prevalent hypertension in fully adjusted models

   e. Table S5. Sensitivity analysis for the association of *HBA* copy number with prevalent hypertension – fully adjusted model with hemoglobin removed

   f. Table S6. Sensitivity analysis for the association of *HBA* copy number with prevalent hypertension – fully adjusted model with chronic kidney disease removed

   g. Table S7. Sensitivity analysis for the association of *HBA* copy number with prevalent hypertension – fully adjusted model with hypertension based on 2017

ACC/AHA guideline (systolic blood pressure ≥ 130 or diastolic blood pressure ≥ 80 mm Hg)

II. Supplemental Methods

a. *HBA* Genotyping Methods

b. Multiple Imputation Procedure

c. Assessment of the Missing at Random Assumption

d. Diagnostic Modeling Description

I. Supplemental Tables

Table S1. Sensitivity analysis for the association of alpha globin gene copy number with prevalent hypertension – fully adjusted model with the addition of the principal components of ancestry*

| | **Prevalent Hypertension**† (n=7,471) | |
|---|---|---|
| | Modified Poisson | |
| | **PR** | **CI** |
| ***HBA* copy number** | 1.01 | (0.98, 1.03) |
| **First 10 PCA**‡ | | |
| **PCA 1** | 1.03 | (1.01, 1.04) |
| **PCA 2** | 0.99 | (0.98, 1.00) |
| **PCA 3** | 1.01 | (0.99, 1.02) |
| **PCA 4** | 1.00 | (0.98, 1.01) |
| **PCA 5** | 1.00 | (0.99, 1.02) |
| **PCA 6** | 1.00 | (0.99, 1.01) |
| **PCA 7** | 1.00 | (0.99, 1.01) |
| **PCA 8** | 1.00 | (0.99, 1.00) |
| **PCA 9** | 0.99 | (0.98, 1.01) |
| **PCA 10** | 1.00 | (0.99, 1.00) |

PCA= principal components of ancestry; CI= confidence interval; $\hat{\beta}$= estimated linear regression coefficient; *HBA*= alpha globin gene copy number.

*The following variables were included in the model but are not displayed in this table: age, sex, body mass index, region, medically insured, education level, income, hemoglobin, kidney disease, diabetes mellitus, total cholesterol, and smoking status. The principal components of ancestry, body mass index, and total cholesterol were scaled by standard deviation. †Prevalent hypertension was defined as one or more of the following: 1) systolic blood pressure $\geq$ 140 mmHg or diastolic blood pressure $\geq$ 90 mmHg; 2) participant self-reported current medication use to control blood pressure; 3) or two or more antihypertensive agents were found on in-home pill bottle review. ‡A subset of the study population (n=7,792 [80%]) had data available for the principal components of ancestry analysis.

Table S2. Sensitivity analysis for the association of alpha globin gene copy number with prevalent hypertension – fully adjusted model with the addition of C-reactive protein*

| | **Prevalent Hypertension**† (n=9,533) | |
|---|---|---|
| | Modified Poisson | |
| | **PR** | **CI** |
| ***HBA* copy number** | 1.00 | (0.98, 1.02) |
| **Log C-reactive Protein** | 1.03 | (1.02, 1.04) |

PR= prevalence ratio; CI= confidence interval; HBA= alpha globin gene copy number.

*The following variables were included in the model but are not displayed in this table: age, sex, body mass index, region, medically insured, education level, income, hemoglobin, kidney disease, diabetes mellitus, total cholesterol, and smoking status. C-reactive protein was evaluated on the log base ten scale and body mass index, and total cholesterol were scaled by standard deviation. †Prevalent hypertension was defined as one or more of the following were true: 1) systolic blood pressure $\geq$ 140 mmHg or diastolic blood pressure $\geq$ 90 mmHg; 2) participant self-reported current medication use to control blood pressure; 3) or two or more antihypertensive agents were found on in-home pill bottle review.

Table S3. Sensitivity analysis for the association of *HBA* copy number with resistant hypertension requiring diuretic use – fully adjusted analysis

| | Resistant Hypertension* (n=9,684) | |
|---|---|---|
| | Modified Poisson | |
| | **PR** | **CI** |
| ***HBA* Copy Number** | 0.96 | (0.86, 1.07) |
| **Age, per year** | 1.02 | (1.01, 1.02) |
| **Sex** | | |
| Female (ref) † | | |
| Male | 1.25 | (1.07, 1.47) |
| **Body mass index‡** | 1.31 | (1.24, 1.39) |
| **Region** | | |
| Non-Belt (ref) | | |
| Belt | 1.02 | (0.88, 1.17) |
| Buckle | 0.98 | (0.82, 1.16) |
| **Medically insured** | | |
| No (ref) | | |
| Yes | 1.07 | (0.84, 1.36) |
| **Education level** | | |
| < HS Grad (ref) | | |
| HS Grad | 1.02 | (0.86, 1.20) |
| Some College | 0.78 | (0.64, 0.94) |
| ≥ College Grad | 0.87 | (0.71, 1.06) |
| **Income** | | |
| < $20K (ref) | | |
| $20K - $34K | 0.92 | (0.78, 1.09) |
| $35K - $74K | 0.96 | (0.79, 1.16) |
| ≥ $75K | 0.70 | (0.51, 1.97) |
| **Hemoglobin, per 1 g/dL** | 0.96 | (0.91, 1.01) |
| **Kidney disease** | | |
| No (ref) | | |
| Yes | 1.94 | (1.69, 2.23) |
| **Diabetes mellitus** | | |
| No (ref) | | |
| Yes | 1.72 | (1.50, 1.97) |
| **Total Cholesterol‡** | 0.85 | (0.79, 0.91) |
| **Smoking status** | | |
| Never (ref) | | |
| Past | 1.03 | (0.90, 1.17) |
| Present | 0.88 | (0.72, 1.08) |

*HBA*= alpha globin gene; PR= prevalence ratio; CI= confidence interval; K= thousand; HS= high school.

*Resistant hypertension was defined in this sensitivity analysis by requiring at least one antihypertensive medication to be a diuretic, and otherwise taking medications from 4 or more antihypertensive classes or systolic blood pressure $\geq$ 140 mmHg or diastolic blood pressure $\geq$ 90 mmHg while taking medications from $\geq$ 3 antihypertensive classes. The median (25$^{th}$, 75$^{th}$ percentile) time elapsed between the baseline and second visit was (9.2 [8.6, 9.9]) years. ‡ref) indicates reference category used for categorized explanatory variables. ‡(Body mass index and total cholesterol were scaled by standard deviation. Multiple imputations were performed for missing data.

Table S4. Pre-specified tests for interaction between *HBA* copy number and age, sex, and chronic kidney disease on prevalent hypertension in fully adjusted models

| | **Prevalent hypertension**<br>Modified Poisson (n=9,684) | | |
|---|---|---|---|
| **Separate fully adjusted models with interaction terms individually added** | **PR** | **CI** | ***P value***[‡] |
| **Age****HBA*** | 1.00 | (1.00, 1.00) | 0.50 |
| **Male Sex****HBA*** | 1.00 | (0.96, 1.03) | 0.89 |
| **Chronic Kidney Disease****HBA*** | 1.01 | (0.97, 1.04) | 0.75 |

*HBA*= alpha globin gene; PR= prevalence ratio; CI= 95% confidence interval

[†]Prevalent hypertension was defined as one or more of the following were true: 1) systolic blood pressure $\geq$ 140 mmHg or diastolic blood pressure $\geq$ 90 mmHg; 2) participant self-reported current medication use to control blood pressure; 3) or two or more antihypertensive agents were found on in-home pill bottle review. [‡]P values calculated by modified Poisson regression model for dichotomous outcome measures

Table S5. Sensitivity analysis for the association of *HBA* copy number with prevalent hypertension – fully adjusted model with hemoglobin removed

| | **Prevalent hypertension*** (n=9,684) Modified Poisson | |
|---|---|---|
| | **PR** | **CI** |
| ***HBA* copy number** | 1.00 | (0.98,1.01) |
| **Age, per year** | 1.01 | (1.01, 1.01) |
| **Sex** | | |
| Female (ref) † | | |
| Male | 1.00 | (0.97, 1.02) |
| **Body mass index‡** | 1.10 | (1.09, 1.11) |
| **Region** | | |
| Non-Belt (ref) | | |
| Belt | 1.05 | (1.02, 1.07) |
| Buckle | 1.02 | (0.99, 1.05) |
| **Medically insured** | | |
| No (ref) | | |
| Yes | 1.07 | (1.02, 1.12) |
| **Education level** | | |
| < HS Grad (ref) | | |
| HS Grad | 1.00 | (0.97, 1.02) |
| Some College | 0.98 | (0.95, 1.01) |
| ≥ College Grad | 0.95 | (0.92, 0.98) |
| **Income** | | |
| < $20K (ref) | | |
| $20K - $34K | 0.99 | (0.96, 1.01) |
| $35K - $74K | 0.96 | (0.93, 0.99) |
| ≥ $75K | 0.94 | (0.90, 0.99) |
| **Chronic Kidney disease** | | |
| No (ref) | | |
| Yes | 1.14 | (1.12, 1.16) |
| **Diabetes mellitus** | | |
| No (ref) | | |
| Yes | 1.16 | (1.14, 1.18) |
| **Total Cholesterol‡** | 0.97 | (0.96, 0.98) |
| **Smoking status** | | |
| Never (ref) | | |
| Past | 1.04 | (1.02, 1.06) |
| Present | 1.04 | (1.01, 1.08) |

*HBA*= alpha globin gene; PR= estimated prevalence ratio; CI= confidence interval; K= thousand; HS= high school.

*Prevalent hypertension was defined as having one or more of the following: 1) systolic blood pressure $\geq$ 140 mmHg or diastolic blood pressure $\geq$ 90 mmHg; 2) self-reported use of medication to control blood pressure; or 3) two or more antihypertensive medications found on in-home pill bottle review; [†](ref) indicates reference category used for categorized explanatory variables. [‡]Body mass index and total cholesterol were scaled by standard deviation. Multiple imputations were performed for missing data.

Table S6. Sensitivity analysis for the association of *HBA* copy number with prevalent hypertension – fully adjusted model with chronic kidney disease removed

| | **Prevalent hypertension*** (n=9,684) Modified Poisson | |
|---|---|---|
| | **PR** | **CI** |
| ***HBA* copy number** | 1.00 | (0.98,1.02) |
| **Age, per year** | 1.01 | (1.01, 1.01) |
| **Sex** | | |
| Female (ref) † | | |
| Male | 1.01 | (0.99, 1.04) |
| **Body mass index‡** | 1.10 | (1.09, 1.11) |
| **Region** | | |
| Non-Belt (ref) | | |
| Belt | 1.05 | (1.02, 1.07) |
| Buckle | 1.02 | (0.99, 1.05) |
| **Medically insured** | | |
| No (ref) | | |
| Yes | 1.07 | (1.02, 1.12) |
| **Education level** | | |
| < HS Grad (ref) | | |
| HS Grad | 0.99 | (0.97, 1.02) |
| Some College | 0.98 | (0.95, 1.01) |
| ≥ College Grad | 0.94 | (0.91, 0.98) |
| **Income** | | |
| < $20K (ref) | | |
| $20K - $34K | 0.98 | (0.96, 1.01) |
| $35K - $74K | 0.96 | (0.93, 0.99) |
| ≥ $75K | 0.94 | (0.89, 0.98) |
| **Hemoglobin, per 1 g/dL** | 0.99 | (0.98, 1.00) |
| **Diabetes mellitus** | | |
| No (ref) | | |
| Yes | 1.19 | (1.16, 1.21) |
| **Total Cholesterol‡** | 0.97 | (0.96, 0.98) |
| **Smoking status** | | |
| Never (ref) | | |
| Past | 1.04 | (1.02, 1.06) |
| Present | 1.05 | (1.02, 1.09) |

*HBA*= alpha globin gene; PR= estimated prevalence ratio; CI= confidence interval; K= thousand; HS= high school.

*Prevalent hypertension was defined as having one or more of the following: 1) systolic blood pressure $\geq$ 140 mmHg or diastolic blood pressure $\geq$ 90 mmHg; 2) self-reported use of medication to control blood pressure; or 3) two or more antihypertensive medications found on in-home pill bottle review; †(ref) indicates reference category used for categorized explanatory variables. ‡Body mass index and total cholesterol were scaled by standard deviation. Multiple imputations were performed for missing data.

Table S7. Sensitivity analysis for the association of *HBA* copy number with prevalent hypertension – fully adjusted model with hypertension based on 2017 ACC/AHA guideline (systolic blood pressure ≥ 130 or diastolic blood pressure ≥ 80 mm Hg)

| | **Prevalent hypertension*** (n=9,684) Modified Poisson | |
|---|---|---|
| | **PR** | **CI** |
| ***HBA* copy number** | 1.00 | (0.99,1.01) |
| **Age, per year** | 1.01 | (1.00, 1.01) |
| **Sex** | | |
| Female (ref) † | | |
| Male | 1.03 | (1.01, 1.05) |
| **Body mass index‡** | 1.07 | (1.07, 1.08) |
| **Region** | | |
| Non-Belt (ref) | | |
| Belt | 1.00 | (0.99, 1.02) |
| Buckle | 1.00 | (0.97, 1.02) |
| **Medically insured** | | |
| No (ref) | | |
| Yes | 1.01 | (0.98, 1.04) |
| **Education level** | | |
| < HS Grad (ref) | | |
| HS Grad | 0.99 | (0.97, 1.01) |
| Some College | 0.99 | (0.96, 1.01) |
| ≥ College Grad | 0.97 | (0.95, 1.00) |
| **Income** | | |
| < $20K (ref) | | |
| $20K - $34K | 0.99 | (0.97, 1.01) |
| $35K - $74K | 0.98 | (0.96, 1.00) |
| ≥ $75K | 0.97 | (0.94, 1.00) |
| **Hemoglobin, per 1 g/dL** | 1.00 | (1.00, 1.01) |
| **Diabetes mellitus** | | |
| No (ref) | | |
| Yes | 1.07 | (1.05, 1.08) |
| **Total Cholesterol‡** | 0.99 | (0.98, 1.00) |
| **Smoking status** | | |
| Never (ref) | | |
| Past | 1.02 | (1.01, 1.04) |
| Present | 1.02 | (1.00, 1.05) |

*HBA*= alpha globin gene; PR= estimated prevalence ratio; CI= confidence interval; K= thousand; HS= high school.

*Prevalent hypertension was defined for this sensitivity analysis as having one or more of the following: 1) systolic blood pressure $\geq$ 130 or diastolic blood pressure $\geq$ 80; 2) self-reported use of medication to control blood pressure; or 3) two or more antihypertensive medications found on in-home pill bottle review; [†](ref) indicates reference category used for categorized explanatory variables. This definition identified 87% (8,410/ 9, 684) participants with prevalent hypertension. [‡]Body mass index and total cholesterol were scaled by standard deviation. Multiple imputations were performed for missing data.

### II. Supplemental Methods

#### a. *HBA* Genotyping Methods

Two-dimensional clusters of droplet counts for target and reference genes were manually gated using Quantasoft (Bio-Rad) per the manufacturer's protocols. Droplet counts, copy number variant (CNV) values, and 95% CIs for CNV were extracted, visualized, and genotype was assigned using custom scripts in the R computing environment without user intervention. A subset of samples was validated against an independent approach employing multiple ligation-dependent probe amplification (MLPA) performed at the Mayo Clinic Laboratory, with 100% concordance. Inter-day variation of our assay was determined by performing the assay on two different days on 672 samples; quantitative copy number varied by less than 1% between days. Reference samples of known genotype were run as positive controls and reaction wells with water instead of DNA were run as negative controls each day.

#### b. Multiple Imputation Procedure

Rather than omit individuals with any missing data from the regression procedures a multiple imputation approach was employed.[13] The analyses reported in Tables 2, 3, and 4 utilized the imputed data. In general, results of the *HBA* copy number effects did not appreciably change with the use of imputed data although estimates for other covariates with significant effects were typically stronger with imputed data. Multiple imputation methods were used in the multivariable analyses. Data on the degree of missingness are described in the footnote to Table 1 in the manuscript. The R package “mice” Version 3.8.0 was used to create and analyze the resulting imputations (Van Buuren S, Groothuis-Oudshoorn K, 2011. mice: Multivariate Imputation by Chained Equations in R, Journal of Statistical Software, 45 (3): 1–67). Each analysis presented is based upon 20 imputations (each developed using 30

Markov Chain based iterations) and the final model coefficients and their standard errors were derived using Rubin's method for pooling results across imputations (Rubin, DB, 1987, Multiple Imputation for Nonresponse in Surveys, John Wiley & Sons, New York, pp. 76-77). The variables used in the imputation procedure were those used in the corresponding regression model. The imputations evolution over 30 iterations was examined visually for convergence and mixing. Further, the distributions of complete and imputed values were visually examined for aberrations.

**c. Assessment of the Missing at Random Assumption**

Missingness was generally rare (<1%) among outcomes and covariates with the exception of hemoglobin (32%) and self-reported income (12%). Hemoglobin is missing primarily because it was not initially collected for approximately the first 8000 of the REGARDS 30239 participants (all races combined). Given the administrative nature of the missing data, an assumption of hemoglobin missing at random (i.e., the probability of missing depends on observed information rather than the underlying missing hemoglobin value) and consequent use of multiple imputation appears reasonable.

Income data reported as missing here reflect refusal to provide information. These self-reported income data may not be missing at random as the refusals might be more likely to coincide with higher or lower than average incomes. As a sensitivity analysis we first imputed the annual income category (either "less than $20k", "$20k-$34k", "$35k-$74k", or "$75k and above") using the multiple imputation algorithm and then moved the imputed category values one level higher if they were not already in the highest category. For example, if a person had an original imputed value of $20k-$34k then in this sensitivity analysis they would now have a value of $35k-$74k. This corresponds to people refusing to answer having higher incomes than predicted. The resulting analysis while the education and income coefficients change marginally, the remaining coefficients and p-values are essentially unchanged from those presented in the prevalence HTN analysis (e.g., the PR estimate and p-value for the HBA copy number are 1.00 and 0.85 - essentially unchanged from the original imputation procedure). The results when lowering (instead of raising) the imputed income category are qualitatively similar with the resulting PR estimate and p-value for HBA copy number 1.00 and 0.85. These results suggest that using a missing at random assumption for income does not likely lead to misleading estimates for any of the covariates. In addition, we note the income categories of "$35k-$74k" and "$75k and above" retained their nominally significant p-values < 0.05 for both the raised and lowered modifications to the original imputation procedure.

**d. Diagnostic Modeling Description**

Our modeling was prespecified in our analytic plan, as described. We performed diagnostic investigation of the Poisson models for HTN prevalence.

For the Poisson models the R function “glm” and R package “sandwich” were used. Residuals were examined for evidence of poor fitting as evidenced by correlation between residuals and predictors of fitted values. Testing of Pearson residuals indicated that age and body mass index were perhaps inadequately modeled as having linear relationships on the log of the risk for HTN prevalence. Consequently, we extended our main model to include quadratic terms for age and hemoglobin. These additional terms had significant p-values but did not change the results for allele count in any meaningful way (point estimates of the risk ratio were unchanged from 1.00 and the p-value changed from 0.85 to 0.95).
